# Alternatives to mental health admissions for children and adolescents experiencing mental health crises: a systematic review of the literature

**DOI:** 10.1101/2021.05.11.21257031

**Authors:** Denisa Clisu, Imogen Layther, Deborah Dover, Russell M. Viner, Tina Read, David Cheesman, Sally Hodges, Lee D Hudson

## Abstract

**Background:** Increasingly more children and young people (CYP) present in mental health crises, many being hospitalised due to concerns around illness severity and lack of community services. To release the burden of admission, we systematically reviewed the literature on the effects of proposed alternatives to CYP in crises.

**Methods:** Three databases (PsychInfo, PubMed and Web of Science) were searched for peer-reviewed papers in October 2020, with an updated search in May 2021.

**Results:** We identified 19 papers of interventions delivered in the emergency department, the home, outside of home but outside of clinics and in hospital clinics. The best evidence came from in-home interventions, in particular multisystemic therapy (MST), which proved to be promising alternatives by improving psychological outcomes and decreasing length of inpatient stay. The quality of included studies was low, with less than half being randomised controlled trials and only half of these at low risk of bias.

**Conclusions:** We could not recommend a particular intervention as an alternative to inpatient admission, however our review describes benefits across a range of types of inteventions that might be considered in multi-modal treatments. We also provide recommendations for future research, in particular the evaluation of new interventions as they emerge.

## Background

Mental health disorders are a substantial burden for children and young people’s (CYP) health globally (Polanczyk, Salum, Sugaya, Caye, & Rohde, 2015), with suicide a leading cause of death (Wasserman, Cheng, & Jiang, 2005). In the UK, recent data has shown that 16% of CYP have a mental health disorder, with over half of older adolescents with a disorder having self-harmed or attempted suicide (Vizard, Sadler, & Ford, 2020). Similar prevalence rates have been reported in Europe and the United States (Kovess-Masfety et al., 2016; Merikangas et al., 2010). Many CYP with mental health disorders will present to health care providers with an acute (sometimes called psychiatric) crisis due to their mental health. Such crises can be defined as subjective experiences where a change in mental wellbeing occurs and the person becomes unstable or at risk to themselves or others (Jennings & Child). In high income countries, numbers of such presentations for CYP seemed to have increased, both to secondary and primary care (Mahajan et al., 2009; Morgan et al., 2017; Newton et al., 2009; Pittsenbarger & Mannix, 2014). Severity of illness, concern about risk (especially in relation to suicide) (Hawton et al., 2012), available community services (Lancet, 2020) and social circumstances (Paranjothy et al., 2018) may mean that such presentations result in an inpatient admission. Whilst hospitalization rates for most paediatric conditions in high income countries have decreased in recent years, admissions because of mental health have increased (Torio, Encinosa, Berdahl, McCormick, & Simpson, 2015).

Inpatient mental health admissions can provide important and vital services for significantly unwell CYP (Green et al., 2007), yet they can also carry substantial burden. Mental health admissions can be lengthy, in locations away from an usual place of residence, leading to disconnection from friends and family, and separation from education or employment. These burdens are especially amplified for CYP experiencing repeated admissions (Miller et al., 2020). Inpatient mental admissions are also more costly for health care systems (Green et al., 2007) versus outpatient care. Demand can also oustrip capacity (O’Herlihy et al., 2003), resulting in admissions of CYP in adult psychiatric wards or non-mental health inpatient settings such as paediatric medical wards (Worrall et al., 2004). Safe and effective interventions acting as alternatives to inpatient admissions for CYP presenting in crisis are therefore highly favourable. Policy makers have turned attention on to this issue, for example in the UK there are strategies in place to improve community services (Alderwick & Dixon, 2019).

Developing and implementing alternatives to inpatient mental health admissions for CYP presenting in crisis requires an up-to-date synthesis of the literature. Previous systematic reviews on this topic (Kwok, Yuan, & Ougrin, 2016; Shepperd et al., 2009) are now outdated (with the latest literature search performed in 2014) and also included papers of interventions with an admission component (such as short-term hospitalisations) which could be a confounder for the effects of proposed alternatives. We therefore systematically reviewed the literature for studies of interventions reported as alternatives to a mental health admission in CYP presenting with a significant mental health crisis. We specifically examined for:

1) effectiveness at avoiding admission or any impact on reducing the length of an inpatient stay if one followed.
2) Improvements in psychological parameters for CYP secondary to such interventions.

## Methods

We searched three databases: PsychInfo, PubMed and Web of Science in October 2020, with an updated search in May 2021. We used search terms to encompass “children and young people”, “mental health crisis” and “potential locations of care” (appendix A). Searches were conducted individually by two researchers (DC and IL) who selected abstracts for inclusion or exclusion. Papers were then downloaded and considered independently, with LH providing adjudication. Reference lists within studies were also screened.

We included studies reporting outcomes of interventions specifically as alternatives to a mental health admission for CYP presenting with a mental health crisis. We defined admission as any hospitalization in any inpatient setting (including general medical settings). We excluded: 1) studies where some or all participants were >18 years; 2) studies not published in English; 3) reviews; 4) papers which did not provide any outcome measures or insufficient outcome measures, or only described interventions; 4) studies where the intervention included an admission.

Independent bias assessments were conducted by DC and IL using the Cochrane Review tools for assessing risk of bias in randomised trials (RCT) (Sterne et al., 2019) and non-RCT (Sterne et al., 2016). Discrepancies were discussed for agreement with final adjudication by LH.

## Results

We found 782 papers in initial searches of databases and were left with 640 unique studies after duplicate removal. 71 papers were retrieved, with 60 excluded based on full text assessment. We found an additional 8 studies from screening reference lists. We included a total of 19 studies. A summary of the search with numbers is shown in figure 1.

**Figure 1.**
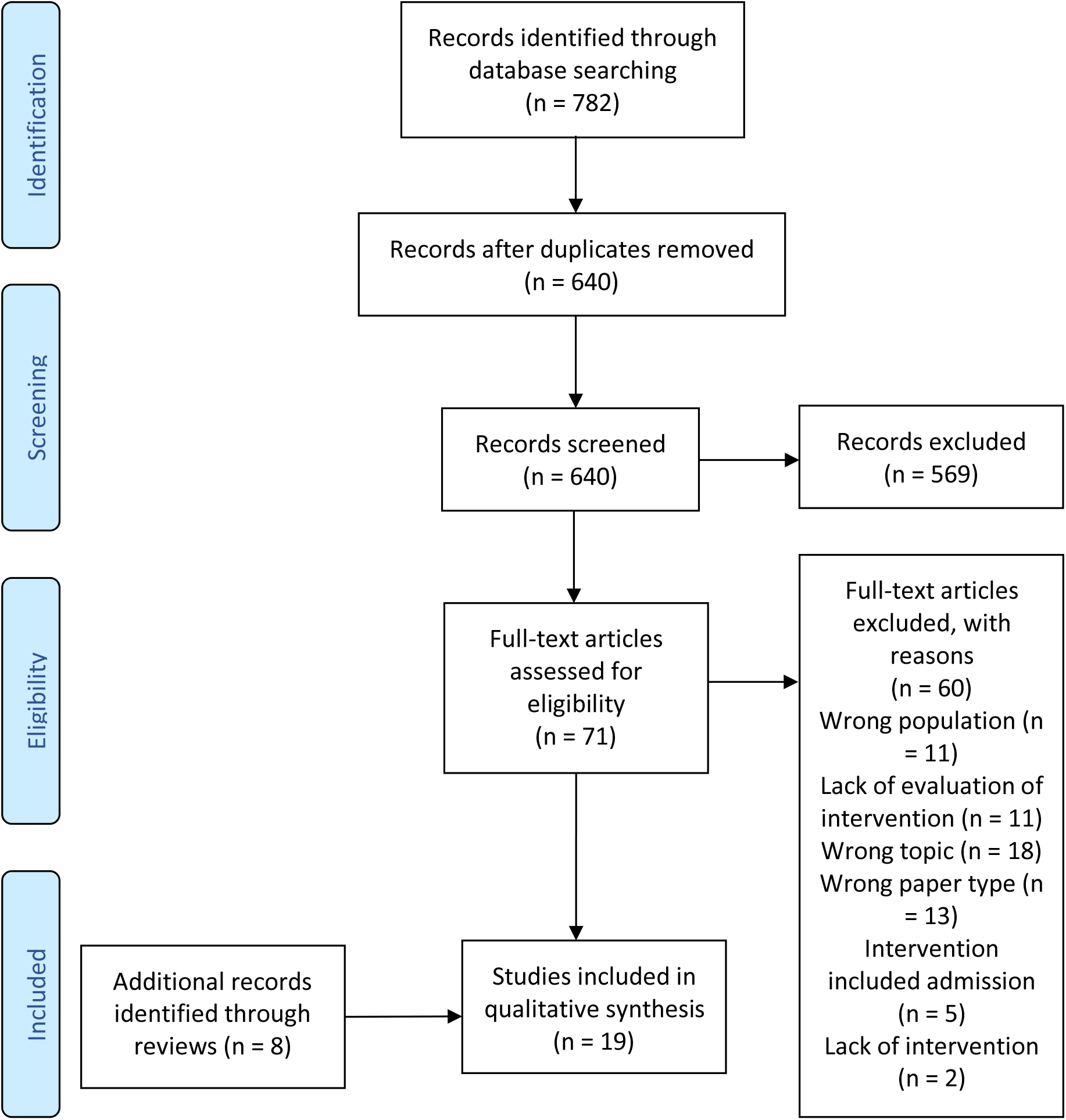
PRISMA flow diagram of included and excluded studies

8 studies were RCTs, 4 studies were service evaluations, 4 studies used an uncontrolled pre-post-treatment investigation, 2 papers used pre-intervention historical/retrospective control groups, and 1 paper used a matched control group. 14 papers were conducted in the USA, 2 papers were conducted in Germany, 2 papers were conducted in Canada and 1 paper was conducted in the UK. 2 studies reported on interventions specifically for CYP with suicidal risk, 1 for psychosis, 1 for disruptive behaviour/externalising crisis presentations and 15 were for mixed types of crisis mental health presentations. For RCTs, we found 4 papers at low risk of bias, 2 raising some concerns and 2 at high risk of bias, and for non-RCTs we found 1 paper at low risk of bias, 2 papers at moderate risk, 6 papers at serious risk of bias and 2 papers at critical risk. Detailed summaries of included studies, including bias assessment are shown in table 1. Detailed rationale for classifying final bias category for each study is shown in Appendix B.

**Table 1.** Summary of the studies included

| Author (year), country | Design | Sample | N (%male, %female) | Age Range (mean, SD) | Outcome Measure(s) | Intervention (duration) | Intervention details | Comparison Condition | Main findings | Conclusions | Overall risk of bias |
| --- | --- | --- | --- | --- | --- | --- | --- | --- | --- | --- | --- |
| Single-session interventions for emergency department crisis presentations |  |  |  |  |  |  |  |  |  |  |  |
| Gillig (2004), USA | Service Evaluation | Adolescents admitted for emergency hospitalisation evaluation | 48 (45.83%, 54.17%) | 12 - 18 (modal age: 16.5) | Inpatient Admission Rate | Emergency Evaluation Interview + brief therapeutic intervention (max. 24 hours) | Semi-structured interview and mental status evaluation. After that, a supportive, reality-based, present-focused therapeutic intervention was offered. Adolescents could be helped for up to 24 hours if necessary. | N/A | 5 patients were hospitalised right after the evaluation (10%). 0 patients were hospitalised in the following month. 4.2% (n = 2) patients were hospitalised within 6 months. | The majority of the adolescents referred to emergency hospitalisation were maintained in the community after receiving the emergency evaluation and brief therapeutic intervention. | Critical |
| Parker et al (2003), Canada | Service Evaluation | Adolescents with psychiatric crisis | n.s. | n.s. | Admission rate and length of inpatient stay | Rapid Response Model (single session offered max 48 hrs post-referral) | Brief interview focused on crisis and risk. | Pre-implementation | Site 1: RRM phase had no significant impact on number of consultations leading to admission. RRM phase had a significant effect on the monthly average length of inpatient stay ($F(1,42) = 3.1, p < .05$): $M_1 = 14.8$, $M_{\text{first RRM}} = 11.2$, $M_{\text{RRM termination}} = 9.5$, $M_{\text{second RRM}} = 19.3$.<br>Site 2: Significant effects of RRM phase on number of consultations leading to admission: decrease from 22% pre-RRM to 2% post-RRM ($\chi^2 = 31.6, N = 340, d.f. = 1, p < .001$). Effect of RRM phase on length of inpatient stay not investigated. | Suggestion of lower admission rate and length of stay associated with RRM implementation, although each was true or investigated only for one site rather than both. | Serious |
| Wharff, Ginnis & Ross (2012) USA | Cohort External Historical Retrospective Control Group | Suicidal adolescents at the ED | In experimental condition: 100 (24%, 76%) | 13 - 18 (15.6, 1.5) | Admission rate | Family-based crisis intervention (FBCI) in the emergency room (single session) | CBT-and family-based session using psychoeducation, therapeutic readiness and safety planning. | Suicidal adolescents (n = 150) at ED before the implementation of the intervention | Significant decrease in admission rate from pre-FBCI (55%) to post-FBCI (35%): $p < .0001$. | FBCI was associated with a lower percentage of patients admitted to hospital in comparison to a control group attending the clinic before its implementation. | Low |

| Exclusively in-home interventions |  |  |  |  |  |  |  |  |  |  |  |
| --- | --- | --- | --- | --- | --- | --- | --- | --- | --- | --- | --- |
| Henggeler et al (1999), USA | RCT | Adolescents referred to emergency psychiatric hospitalisation | 113 (65%, 35%) | 10-17 (13, n.s.) | GSI-BSI, CBCL, PEI, Self-Esteem subscale of FFS | Multisystemic Therapy (3 - 6 months) | Family and behavioural therapy strategies used to intervene in the key social systems. | Hospitalisation (n = 56) | Both groups reported pre- to post-treatment improvements in GSI-BSI (child and caregiver reported), CBCL internalising (caregiver reported) and CBCL Social functioning (caregiver reported), and a deterioration in FACES-III Family Adaptability (caregiver) ( $p < .05$ ). MST was reported to be superior to hospitalisation in improving externalising symptoms in both caregiver and teacher reports: $F(1,102) = 6.55, p < .011$ and $F(1, 45) = 4.10, p < .048$ . MST patients reported a pre- to post-treatment decrease from $M = 73.3$ ( $SD = 10.3$ ) to $M = 63.7$ ( $SD = 12.4$ ) in caregiver-reports and $M = 71.1$ ( $SD = 10.7$ ) to $M = 64.8$ ( $SD = 11.8$ ) in teacher-reports. Hospitalised patients reported a pre- post-treatment decrease from $M = 70.6$ ( $SD = 12.3$ ) to $M = 64.3$ ( $SD = 14.2$ ) in caregiver reports and an increase from $M = 67.8$ ( $SD = 15.1$ ) to $M = 68.0$ ( $SD = 13.0$ ) in teacher reports. MST was also reported to be superior than hospitalisation in FACES-III family Adaptability (youth-reported) and Faces-III family Cohesion (caregiver-reported): $F(2, 220) = 3.28, p = .039$ and $F(2, 206) = 6.56, p < .001$ . Families in the MST condition became more<br>structured M = 23.1 (SD = 6.7) to M = 21.8 (SD = 8.1) in comparison to admitted patients: M = 22.1 (SD = 6.7) to M = 23.8 (SD = 7.4). In the MST, family cohesion increased from pre- to post-treatment from M = 32.2 (SD = 8.4) to M = 34.4 (SD = 6.6) while this decreased in the hospitalisation condition from M = 36.1 (SD = 5.3) to M = 34.7 (SD = 6.4). Patients in the MST condition spent less day out of school (FFS) by the end-of-treatment (M = 14 SD = 36.8) than hospitalised patients (M = 37 SD = 59.8): $F(1,110) = 5.72$, $p < .001$. Hospitalisation was reported to be superior to MST in increasing youth self-esteem (FFS): $F(1, 109) = 7.72$, $p = .006$. Hospitalised patients increased their pre- post-treatment score from M = 2.21 (SD = 1.0) to M = 2.73 (SD = 0.9) while patients in the MST condition decreased their pre- post-treatment score from M = 2.57 (SD = 0.9) to M = 2.55 (SD = 1.1). No significant time- or group-differences were reported for PEI alcohol, marijuana and arrests, Faces-III Family Cohesion (youth-reported), CBCL-social functioning (youth-reported), FFS Conventional Involvement<br>and FFS Antisocial Friend (p >.05). | While both treatments improved child's self-reported and caregiver-reported symptoms severity, caregiver-reported internalising symptoms and social functioning and family adaptability as reported by youths, MST proved to be superior to hospitalisation in many areas. For example, MST was better than hospitalisation at improving externalising symptoms (caregiver and teacher reports), family adaptability (youth<br>reported) and cohesion (caregiver reported) and was associated with less days spent out of school. Hospitalisation was however superior to MST in improving youth self-esteem. | Low |
| Henggeler et al (2003), USA | RCT | Adolescents referred to emergency psychiatric hospitalisation | 113 (65%, 35%) | 10-17 (12.9, 2.1) | GSI-BSI, CBCL, PEI, Self-Esteem subscale of FFS | Multisystemic Therapy (3 - 6 months) | Family and behavioural therapy strategies used to intervene in the key social systems. | Hospitalisation (n = 56) | Clinical improvements were maintained until the 12-months follow up point, but the group differences which were initially found in Henggeler et al (1999) and Huey et al (2003) dissipated. | Both MST and hospitalisation were effective at improving the of CYP in crises on the long term, but initial differences seen between groups dissipated at one-year post-treatment. | Low |
| Huey et al (2003), USA | RCT | Adolescents referred to emergency psychiatric hospitalisation | 113 (65%, 35%) | 10-17 (12.9, 2.1) | FFS, BSI, CBCL, HSC, YRBSS | Multisystemic Therapy (3 - 6 months) | Family and behavioural therapy strategies used to intervene in the key social systems. | Hospitalisation (n = 56) | A significant time effect was found in both groups for most measures, with patients scoring lower at follow-up than pre-treatment in CBCL caregiver-rated attempted suicide (t = -5.25, p < .001), CBCL anxiety/depression (t = -5.70, p < .001), and youth-rated BSI suicidal ideation (t = -5.97, p < .001), BSI depression (t = -3.47, p < .001) and hopelessness (t = -2.65, p < .01). Group differences for these measures were not significant. Patients in the MST condition reported a steeper decrease in child-reported suicide attempts from pre-treatment (31%) to follow-up (4%) than hospitalised CYP (from 19% to 4%): t = 3.60, p < .001. | Compared to hospitalisation, MST was found to be more effective in reducing youth-reported suicide attempts. Both treatments were equally effective in decreasing caregiver-reported attempted suicide, suicidal ideation, hopelessness | Low |
|  |  |  |  |  |  |  |  |  | This effect was not reported for the caregiver-rated attempted suicide where results did not reach statistical significance ($p > .05$). Different trajectories were reported in the two groups for FFS caregiver rated parental control ($t = 2.08$, $p < .05$), with MST parents reporting an initial increase in control at post-treatment ($M = 3.06$, $SD = 0.54$) in comparison to pre-treatment ($M = 2.85$, $SD = 0.58$, but which then decreased back to baseline levels at 1-year follow-up ($M = 2.87$, $SD = 0.59$). In the hospitalisation group, parental control was maintained at similar levels from pre-treatment ($M = 2.92$, $SD = 0.62$) to post-treatment ($M = 2.95$, $SD = 0.51$) and follow-up ($M = 2.92$, $SD = 0.54$). No significant time or group effects were reported for the youth rated FFS parental control. | and depression and anxiety symptoms. |  |
| Lay, Blanz & Schmidt (2001), Germany | Uncontrolled pre-post study | CYP with externalising psychiatric disorders requiring inpatient admission | 50 (74%, 26%) | 5 - 16 (9.8, 2.4) | SGKJ, MEI, Non-involved clinician in the care of the child ratings on: symptom load, level of functioning, psychosocial environment | Home-treatment (1-2/week for 2hrs for 3.5 months) | Utilised principles of behaviour modification and parent training: reinforcement schedules, training of social skills and therapeutic exercises. | N/A | A significant decrease was reported in the MEI total symptoms score from pre-treatment (M = 11.9) to post-treatment (M = 8.1): $p < .001$. Before treatment, $n = 18$ (36%) of the sample scored $\leq 9$, $n = 27$ (54%) scored 10 - 19 and $n = 5$ (10%) scored $\geq 20$ in the MEI. After treatment, $n = 34$ (68%) scored $\leq 9$, $n = 14$ (28%) scored 10 - 19 and $n = 2$ (4%) scored $\geq 20$. This difference between pre- and post-treatment reached statistical significance ($p < .001$). Significant pre- to post-treatment differences were reported in the child-rated psychosocial functioning as well ($p < .001$), with $n = 27$ (54%) and $n = 23$ (46%) scoring 5 and 4 respectively, while post treatment $n = 2$ (4%) scored 4, $n = 20$ (40%) scored 5, $n = 17$ (34%) scored 6, $n = 9$ (18%) scored 7, $n = 1$ (2%) scored 8 and $n = 1$ (2%) scored 9. In comparison to pre-treatment, significantly more participants achieved higher scores in parent-rated psychosocial functioning at post treatment in the following sub-domains: family ($p = .001$), peers ($p = .02$), interests ($p = .001$) and autonomy ($p = .02$), while school performance did not reach statistical significance ($p = .22$) | The home treatment was associated with a decrease in psychological symptoms and psychosocial adjustment, showing positive results in almost all areas of functioning considered. | Serious |
|  |  |  |  |  |  |  |  |  | and school discipline and social behaviour had no data. In comparison to pre-treatment, significantly more participants achieved higher scores in all the therapist-rated sub domains: family (p = .001), peers (p = .002), interests (p = .001), autonomy (p = .001), school performance (p = .004), school discipline (p = .002) and school social behaviour (p = .003). |  |  |
| Mosier, Burlingame & Wells (2001), USA | Uncontrolled pre-post study | CYP who require restrictive and intensive mental health services | 104 (62%, 38%) | 4 - 17 (n.s.) | Y-OQ | In-home family centered treatment | 24/7 access to services, exploring areas to improve as a family system | N/A | Scores on the Y-OQ decreased from pre-treatment through the intervention: pre-treatment M = 106.53 SD = 36.68, time 2 (M = 90.30 SD = 37.26), time 3 (M = 79.48 SD = 40.52), post-treatment (M = 75.17 SD = 37.79). At time 2 (approximately 2 weeks post-referral), out of the 64 people who provided data, 5 recovered (8%), 29 achieved reliable improvements (45%), 30 remained unchanged (47%) and 0 deteriorated (0%). At time 3 (approximately 2 weeks post time 2), out of the 46 people who provided data, 10 patients recovered (22%), 18 reliably improved (39%), 8 remained unchanged (17%) and 10 deteriorated (22%). At time 4 (end-of-treatment), out of the 29 people who provided data, 5 patients recovered (17%), 13 patients improved reliably | The in-home family treatment was suggested to lead to improvements or even recovery for a number of patients, from pre-treatment to throughout the treatment and finalisation. When compared to a group of patients receiving outpatient treatment, no evidence was found for superiority of | Serious |
|  |  |  |  |  |  |  |  |  | (45%), 5 patients remained unchanged (17%) and 6 patients deteriorated (21%). Comparing the mean Y-OQ score at the end of treatment of the in-home family treatment to the normative scores of a sample receiving inpatient/outpatient treatment revealed that although the mean was lower for the in-home intervention (M = 75.17 SD = 37.79) than for the control (M = 78.6 SD = 36.40), this difference did not reach statistical significance (t(369) = 1.17, p > .05). | the in-home intervention. |  |
| Rowland et al (2005), USA | RCT | CYP at risk of out-of-home placement | 31 (58%, 42%) | n.s. (14.5, n.s.) | Out-of-home placement (including admission), CBCL, YRBSS, PEI, Criminal Activity, SRD, school placement, FACES III, SSQ | Multisystemic Therapy (3 - 6 months) | Family and behavioural therapy strategies used to intervene in the key social systems. | Usual Care | A significantly higher change was reported in externalising symptoms for MST in comparison to usual care (F(1, 25) = 4.62, p = .041), with MST children reporting a decrease in scores from M = 66.80 (SD = 12.74) at pre-treatment to M = 60.53 (SD = 13.58) at follow-up and children in usual care reporting similar scores between pre-treatment (M = 63.36, SD = 10.93) and follow-up (M = 63.00, SD = 11.39). A significantly higher change was reported in internalising symptoms for MST in comparison to usual care (F(1,28) = 6.05, p = .021), with MST children reporting a decrease in scores from M = | MST was reported to be superior to usual care in improving youth-rated externalising and internalising symptoms, self-reported minor delinquencies, days spent in general education and monthly days in out of home placement by the end of treatment. | Some concerns |
|  |  |  |  |  |  |  |  |  | 62.27 (SD = 9.79) at pre-treatment to M = 57.07 (SD = 13.19) at follow-up and children in usual care reporting an increase in scores from pre-treatment (M = 57.29, SD = 14.42) to follow-up (M = 59.00, SD = 11.80). MST was also superior in decreasing minor delinquencies in comparison to usual care ($F(1, 25) = 5.74, p = .02$), with MST patients reporting a steeper decrease in scores from M = 26.33 (SD = 26.44) at pre-treatment to M = 5.27 (SD = 8.25) at follow-up, than usual care patients (pre-treatment: M = 5.53 SD = 10.88; follow-up M = 3.13, SD = 5.26). By the end of the trial, youth in the MST condition spent 42% more days per month in general education settings (M = 85.64, SD = 78.91) compared to youth in the usual care condition (M = 49.64, SD = 77.25): $F(1, 24) = 3.16, p = .07$. By the end of the trial, MST patients spent significantly less monthly days in out-of-home placement (M = 3.75 SD = 4.77) than youth in the usual care condition (M = 11.83 SD = 11.46): $F(1, 26) = 5.68, p = .025$. No significant group or time differences were found for youth dangerousness, caregiver-reported internalising and<br>externalising symptoms, substance abuse, self-reported index offenses, arrests/month in the community, family adaptability and cohesion. |  |  |
| Schoenwald et al (2000), USA | RCT | Adolescents referred to emergency psychiatric hospitalisation | 113 (65%, 35%) | 10 - 17 (13, n.s.) | Admission rate and length of stay | Multisystemic Therapy (3 - 6 months) | Family and behavioural therapy strategies used to intervene in the key social systems. | Hospitalisation (n = 56) | 44% of the patients receiving MST were admitted to hospital by the end of treatment. The mean days hospitalised was significantly lower for patients in the MST group (M = 2.39, SD = 4.55) than patients in the control group (M = 8.82, SD = 11.55): $t = 3.91$, $p = .001$. The mean hospitalisation days per hospitalized youth was lower in the MST condition (M = 5.44, SD = 5.58) than the control group (M = 8.82, SD = 11.55), but this did not reach statistical significance: $p > .50$. Patients admitted from MST group spent less days hospitalised per episode (M = 3.78, SD = 5.04) than patients in the control condition (M = 6.06, SD = 4.05), but this difference did not reach statistical significance ($p > .50$). | The majority of patients receiving MST avoided admission by the end of treatment. It was suggested that MST was superior at decreasing the number of overall days hospitalised and days hospitalised per episode in comparison to admission. | Low |
| Schmidt, Lay, Gopel, Naab, & Blanz (2006), Germany | Matched sample control group | CYP requiring psychiatric hospitalization | 105 (n.s.) | 6 - 17 (n.s.) | MEI, Behaviour Changes (child, parents, therapist) | Home treatment (3 months) | Child-centred tailored, home-based and family-focused treatment. | Hospitalisation | Both groups achieved significant positive pre-treatment to follow-up changes in the total MEI symptom score ( $p < .001$ ). Hospitalisation was superior in improving the total symptom score and achieving higher behaviour changes. For home-treatment patients, the MEI<br>symptoms scores decreased from M = 12.0 (SD = 5.2) at pre-treatment to M = 8.0 (SD = 5.2) at post-treatment, while for CYP admitted, the scores decreased from M = 14.8 (SD = 5.4) at pre-treatment to M = 6.2 (SD = 3.7) at post-treatment $p < .001$. Receiving the home treatment was associated with significant improvements from intake to follow-up in 4 of the 5 functioning subscales (family, peers, interests and autonomy) while for hospitalisation, change from pre-treatment to follow-up was only significant in 3 subscales (family, interests, autonomy). When comparing the degree of change from pre-treatment to follow-up between the two treatment groups, no significant differences were reported For the child- and parent-rated behaviour changes at post-treatment, patients in the home-treatment group scored significantly lower (M = 4.1; SD = 0.9/ M = 4.2; SD = 0.7) than patients in the hospitalisation group (M = 4.6; SD = 0.6/ M = 4.5; SD = 0.6): child ($p = .02$), parent ($p = .03$). These changes were not significant ($p = .05$) when comparing the therapist ratings for the two treatment groups. When<br>investigating the blind examiners' rating of each treatment's effectiveness, hospitalisation proved to be superior in every measure: symptoms (hospitalisation: M = 2.2, SD = 0.9; home: M = 1.8, SD = 1.0; p = .03), level of functioning (hospitalisation: M = 2.0, SD = 0.9; home: M = 1.5, SD = 1.0; p = .01), psychosocial environment (hospitalisation: M = 1.9, SD = 0.9; home: M = 1.3, SD = 1.0; p = .008), global rating (hospitalisation: M = 2.1, SD = 0.9; home: M = 1.6, SD = 1.0; p = .01) | CYP admitted had higher significant improvements reported in the total symptoms score ( $p < .001$ ) and child- and<br>parent-rated behaviour Although both treatments led to clinical and social functioning improvements , hospitalisation was superior at improving psychological symptoms and level of functioning and determining positive behaviour changes by the end of treatment. | Moderate |
| Wilmshurt (2002), USA | RCT | CYP with emotional and behavioural disorders (EBD) | 65 (n.s., n.s.) | 6 - 14 (n.s., n.s.) | SCIS, SSRS | Family Preservation Program (up to 12hrs/week, 12 weeks) | Flexible and intensive in-home support available using problem-solving and CBT-based techniques. | 5-day residential program (5DR) (5 days/week, 3 months). Individualised and flexible day treatment. | A significantly higher decrease in internalising symptoms in the family-therapy group was detected from pre- treatment (M = 69.76; SD = 13.3) to 1-year follow (M = 62.58; SD = 11.6) in comparison to the residential treatment where an increase occurred from pre-treatment (M = 65.74; SD = 11.8) to 1 year follow-up (M = 66.41; SD = 12.8): F(2, 62) = 3.92, p = .025. All patients significantly improved in externalising symptoms (F(2, 62) = 28.67, p = .001), behavioural problems (F(2, 66) = 24.89, p = .001) and social competence F (2, 66) = 11.61, p = .001) from pre-treatment to post- treatment and 1 year follow-up, but no significant group differences were reported for these measures (p > .05). | Although both treatments improved externalising and behavioural problems and social competence, the family programme was superior in improving internalising symptoms in comparison to the residential treatment, which was reported to worsen the problems. | Some concerns |
| Outside of the home but outside of hospital clinics |  |  |  |  |  |  |  |  |  |  |  |
| Darwish, Salmon, Ahuja, & Steed (2006), UK | Service Evaluation | CYP with complex psychological needs | 42 (n.s.) | n.s. | Admission rate | Community Intensive Therapy (CITT) (n.s.) | Services offered were similar to inpatient setting but delivered in the community . Strong emphasis on interagency network and especially with educational services. Individual and family therapy were offered flexibly, limited group therapy activities, and dieticians, and clinical psychologists were available if needed. | A similar sample attending the service before the implementation of the CITT | Number of inpatient admissions decreased considerably after CITT implementation: 5-6 CYP were admitted the year before implementation at any one time while for the 5 years after implementation, 0-1 CYP were admitted every year. | CITT was suggested to be effective in reducing the likelihood of inpatient psychiatric admission of CYP with complex mental health needs. | Serious |
| Vanderploeg, Lu, Marshall & Stevens (2016), USA | Service Evaluation | CYP in psychiatric crisis | n.s. | n.s. | The Ohio Scale | Emergency Mobile Psychiatric Services (EMPS) (average of 20.8 days of help, maximum 45 days) | Emergency crises service delivered at various locations 24/7: crisis stabilisation and support, screening and assessment, suicide assessment and prevention, brief solution-focused interventions, and referral and linkage to ongoing care. | N/A | Significant pre- to post-treatment changes occurred in parent- and clinician-rated child functioning and problem severity. Parent-rated functioning scores increased from M = 42.94 at pre-treatment to M = 45.52 at post-treatment (t = 4.70, p < .001), clinician rated functioning scores increased from M = 43.44 at pre-treatment to M = 45.38 at discharge (t = 14.96, p < .001), parent-rated problem severity decreased from M = 28.66 at pre-treatment to M = 23.04 at discharge ( $t = -8.53, p < .001$ ) and clinician rated problem severity decreased from M = 28.51 at intake to M = 25.56 at discharge ( $t = -21.24, p < .001$ ). The percentage of clinically significant change in scores for each measures was reported as follow: 13% for parent-rated functioning, 8.4% for clinician-rated functioning, 19.1% for parent-rated problem severity and 10.4% for clinician-rated problem severity. | EMPS improved psychological outcomes and social functioning and led to clinically significant results for some children. | Critical |
| Winsberg et al. (1980), USA | RCT | CYP with severe behaviour disorders referred to admission | 49 (84%, 16%) | 5 - 13 (n.s.) | BRS, DESB, DCB, MAT, SESAT, PSS (in mothers), FFC | Community Treatment (6 months) | Highlight on support for the parents and involvement of social services for the family. Distinguishing characteristic of the program: continuous availability of staff, persistent advocacy and treatment flexibility. Pharmacol | Inpatient Hospitalisation (6 months) | A significant decrease in scores for all measures was detected for all factors in the Community Treatment Group. A decrease from M = 2.40 (SD = .69) to M = 1.95 (SD = .81) was reported on the BRS aggressivity scale ($t = 2.58, p < .05$), from M = 2.61 (SD = .75) to M = 2.07 (SD = .71) on the BRS inattentiveness scale ($t = 3.53, p < .01$), and from M = 3.18 (SD = .72) to M = 2.35 (SD = .78) on the BRS hyperactivity scale ($t = 4.27, p < .01$). A decrease from M = .85 (SD = .69) to M = .48 (SD = .83) was detected on the DESB ($t = 2.58, p < .05$) and a decrease from M = 1.93 (SD = 1.26) to M = 1.32 (SD = 1.24) was reported on the DCB ($t = 2.22, p < .05$). For the hospitalisation group, pre- to post-treatment differences were | Community treatment was reported to improve more psychological symptoms than hospitalisation, however no statistical tests were conducted to investigate the difference in change between the two groups. Both groups reported similar achievements in arithmetic | High |
|  |  |  |  |  |  |  | ogy or individual therapy provided where needed. |  | not significant on any scale, apart from BRS inattentiveness scale where the scores decreased from M = 1.81 (SD = .69) to M = 1.49 (SD = .41) (t = 2.32 p < .05). The time x treatment group interaction on these measures was not investigated. Patients from both groups reported significant improvements in reading and arithmetic by end of treatment in comparison to intake (p < .05), but the gain ratio (the change between the pre- and post-treatment scores divided by number of school months attended) did not differ significantly between the two treatment groups (p > .05). No significant time or time x group differences were reported for mother's mental health on the PSS or the FFC domains. | and reading by the end of treatment and parental mental health was not impacted (neither positively nor negatively) by either of the two treatments. |  |
| Exclusively clinic-based interventions, including day treatment |  |  |  |  |  |  |  |  |  |  |  |
| Asarnow, Berk, Hughes & Anderson (2015), USA | Uncontrolled pre-post study | Adolescent suicide attempters | 35 (14%, 86%) | n.s. (14.89, 1.60) | NIMH DISC-IV, HASS, CES-D, BHS, SAS | SAFETY Program (M total sessions = 10.14) | Based on cognitive-behavioural techniques, aims to enhance safety and reduce suicide risk. Included individual, parental<br>and family<br>sessions | N/A | Significant pre-treatment to follow-up differences were reported on all scales. HASS suicide attempt scores decreased from M = 0.89 (SD = 1.86) to M = 0.13 (SD = 0.34) (t = 2.42, p = .019, d = .64); HASS active suicide behaviour and ideation decreased from M = 3.71 (SD = 4.42) to M = 1.81 (SD = 2.69) (t = 2.63, p = .019, d = .59); Scores on the HASS passive<br>suicide ideation decreased from M = 12.69 (SD = 9.79) to M = 9.19 (SD = 10.14) ($t = 2.56$, $p = .016$, $d = .39$); total HASS scores decreased from M = 16.40 (SD = 13.52) to M = 11.04 (SD = 12.05) ($t = 2.70$, $p = .011$, $d = .46$); CES-D youth scores decreased from M = 24.54 (SD = 12.33) to M = 13.69 (SD = 9.83) ($t = 4.53$, $p < .001$, $d = .91$); CES-D parent scores decreased from M = 20.26 (SD = 13.34) to M = 10.86 (SD = 8.55) ($t = 3.47$, $p = .002$, $d = .71$); BHS scores decreased from M = 8.86 (SD = 5.70) to M = 3.94 (SD = 3.79) ($t = 5.58$, $p < .001$, $d = 1.01$); SAS total scores decreased from M = 41.96 (SD = 7.07) to M = 32.53 (SD = 7.26) ($t = 6.13$, $p < .001$, $d = 1.27$); SAS school scores decreased from M = 13.90 (SD = 4.94) to M = 10.37 (SD = 3.57) ($t = 3.53$, $p = .002$, $d = .90$); SAS peer scores decreased from M = 17.26 (SD = 4.60) to M = 12.78 (SD = 3.54) ($t = 5.36$, $p < .001$, $d = 1.11$); SAS family scores decreased from M = 14.02 (SD = 3.61) to M = 11.45 (SD = 3.83) ($t = 2.79$, $p = .009$, $d = .66$); SAS spare time scores decreased from M = 5.15 (SD = 1.76) to M = 4.58 (SD = 1.57) ($t = 2.76$, $p = .01$, $d = .52$); | The SAFET program was reported to be effective in improving all clinical, functioning and family outcomes. | Moderate |
| Greenfield<br>, Hechtman<br>, & Tremblay<br>(1995),<br>Canada | Cohort<br>External<br>Historical<br>Retrospective<br>Control<br>Group | CYP in crisis<br>referred from<br>the emergency<br>department | 980 (n.s.) | n.s. | Admission<br>rate | Emergency<br>Room<br>Follow-up<br>Team<br>(ERFUT) | Immediate,<br>intensive<br>(daily if<br>needed),<br>short-term,<br>family and<br>psycho-<br>dynamicall<br>y oriented<br>treatment<br>in addition<br>to other<br>interventio<br>ns | CYP<br>presenting<br>to the<br>emergency<br>room in<br>crisis<br>before the<br>implement<br>ation of the<br>service | A 16% reduction in the<br>proportion of patients admitted<br>was recorded after the<br>implementation of the ERFUT in<br>comparison to a period before<br>its implementation: $p < .001$.<br>Less patients were hospitalised<br>after the implementation ($n = 118$, 21%), in comparison to<br>before ($n = 152$, 37%). A<br>decrease was also reported on<br>the number of people<br>subsequently returning to the<br>emergency room after the<br>impmenentation of the ERFUT ($n = 16$, 2.82%) in comparison to<br>before ($n = 19$, 4.61%), but this<br>difference did not reach<br>statistical significance. No<br>statistical differences were<br>reported between the two study<br>groups in the mean number of<br>return emergency room visits<br>per patient and the number of<br>hospitalizations per patient after<br>a second emergency room visit.<br>No difference was reported in<br>the number of return visits to<br>the emergency room between<br>the patients referred to the<br>ERFUT and the patients who<br>attended the emergency room<br>but were not referred to the<br>ERFUT in the same period. | In comparison<br>to a period<br>before its<br>implementatio<br>n, ERFUT was<br>associated<br>with a<br>decrease in<br>admissions for<br>CYP presenting<br>to the<br>emergency<br>deptartment in<br>psychological<br>crisis. | Serious |
| Silberstein , Mandell, Dalack, Cooper & Island (1967), USA | RCT | Psychotic children and adolescents that require psychiatric hospitalisation | 48 (69%, 31%)) | 4.16 - 17 (10.33, n.s.) | Rate of inpatient admission, hospitalisation requests, police difficulties, out of regular school placements | Parental counselling with or without medication (1 hr/week) | n.s. | No counselling with or without medication | Most of the patients were maintained in the community and no significant differences were reported between the 4 groups in regard to the number of patients hospitalised ($p > .05$). The number of patients who successfully avoided hospitalisation in each group were as follows: parental counselling + medication group - $n = 11$ ($N = 12$, 91.7%); parental counselling + placebo - $n = 11$ ($N = 12$, 91.7%); no counselling + medication - $n = 14$ ($N = 14$, 100%); no counselling + placebo - $n = 10$ ($N = 10$, 100%). No significant group differences occurred for amount of patients who did not provoke hospitalisation requests, did not get into police difficulties and remained in their regular classroom: parental counselling + medication ($n = 11$, $n = 9$, $n = 8$ out of $N = 12$), parental counselling + placebo ($n = 11$, $n = 10$, $n = 9$ out of $N = 12$), no counselling + medication ($n = 13$, $n = 14$, $n = 11$ out of $N = 14$), no counselling + placebo ($n = 10$, $n = 10$, $n = 8$ out of $N = 10$). | The majority of the patients in the trial were maintained in the community, did not provoke hospitalisation requests, did not get into police difficulties and remained in their regular classroom. Neither parental counselling nor medication proved to be more effective in comparison to each other or in comparison to no treatment. | High |
| Kiser et al (1996) USA | Uncontrolled pre-post | CYP moderately to severely affected by a diagnosable psychiatric disorder | 114 (n.s.) | 5 - 18 (11.6, n.s.) | CBCL, YRS, Utilization of Mental Health Services, Intake and follow-up questionnaire (parent report of child functioning), FAD | Partial Hospitalization (12-397 days, M = 115.9) | Individual therapy, family therapy, group psychotherapy, psychoeducational groups | N/A | Treatment significantly decreased scores on all CBCL subscales from intake to follow-up apart from sex problems (from M = 59.36 to M = 55.92, t = 1.33, p = .195); withdrawn (from M = 63.72 to M = 57.95, t = 5.25, p < .001), somatic complaints (from M = 61.47 to M = 58.08, t = 3.11, p = .003); anxious/depressed (from M = 65.81 to M = 61.02, t = 3.95, p < .001); social problems (from M = 65.29 to M = 62.69, t = 2.70, p = .008); thought problems (from M = 65.44 to M = 59.67, t = 5.66, p < .001); attention problems (from M = 69.26 to M = 64.29, t = 4.28, p < .001); delinquent problems (from M = 67.01 to M = 64.12, t = 2.49, p = .015); aggressive behaviour (from M = 70.26 to M = 64.35, t = 4.49, p < .001); total problems (from M = 70.08 to M = 63.14, t = 6.24, p < .001); internalizing (from M = 65.16 to M = 58.59, t = 4.90, p < .001); externalizing (from M = 68.82 to M = 63.34, t = 4.84, p < .001). Treatment significantly decreased scores on all YSR subscales from intake to follow-up: withdrawn (from M = 59.62 to M = 55.80, t = 2.91, p = .005), somatic complaints (from M = 61.02 to M = 56.60, t = 2.76, p = .008); anxious/depressed (from | The partial hospitalisation was successful in improving all clinical outcomes (apart from sex problems as rated by the child) and some school, peer relations and community integration functioning outcomes. Both parents and children reported that affective involvement and behaviour control improved in the family by end of treatment. The treatment also decreased the use of mental health inpatient services. | Critical |
|  |  |  |  |  |  |  |  |  | M = 61.34 to M = 55.94, t = 3.95, p < .001); social problems (from M = 59.76 to M = 55.76, t = 2.38, p = .021); thought problems (from M = 60.36 to M = 55.46, t = 3.28, p = .002); attention problems (from M = 61.50 to M = 56.54, t = 2.89, p = .006); delinquent problems (from M = 64.06 to M = 59.60, t = 3.77, p < .001); aggressive behaviour (from M = 62.58 to M = 57.56, t = 3.59, p < .001); sex problems (from M = 62.83 to M = 54.80, t = 4.66, p < .001); total problems (from M = 63.44 to M = 54.04, t = 5.58, p < .001); internalizing (from M = 60.30 to M = 51.86, t = 4.94, p < .001); externalizing (from M = 63.44 to M = 56.10, t = 4.90, p < .001). Treatment significantly improved parent-rated child functioning levels from intake to follow-up in the following domains: school suspensions (from 39% to 36%, $\chi^2 = 8.78$, df = 1, p < .01), child/adolescents is a good-quality friend (from 65% to 88%, $\chi^2 = 6.45$, df = 1, p < .05), incarcerated (from 4.7% to 6.3%, $\chi^2 = 19.6$, df = 1, p < .01), trouble with police (from 13.4% to 16.4%, $\chi^2 = 11.6$, df = 1, p < .01). The following parent-rated child functioning dimensions did not reach statistical significance<br>from intake to follow-up: school (grade C or above, students failing, conduct: satisfactory or excellent); peer relations (well-adjusted/somewhat adjusted socially, many friends); community integration (on probation while for the eligible for work/holding jobs, no statistical test was conducted). For the parent-rated FAD, treatment was associated with a significant change from intake to follow-up in the following subscales: roles (from M = 2.50 to M = 2.38, t = 2.68, p = .009), behaviour control (from M = 1.88 to M = 1.73, t = 3.41, p < .001) and a marginally significant effect in affective involvement (from M = 2.34 to M = 2.22, t = 1.98, p = .051). The intake-follow-up difference did not reach significance in: problem solving, communication, affective responsiveness, general functioning. For the child rated FAD, treatment was associated with a significant change from intake to follow-up in the following subscales: affective involvement (from M = 2.60 to M = 2.43, t = 2.24, p = .030) and behaviour control (from M = 2.18 to M = 1.92, t = 3.48, p < .001). The intake-follow-up difference did not<br>reach statistical significance in: problem solving, communication, roles, affective responsiveness and general functioning. Use of inpatient treatment decreased from 35.1% at intake to 10.7% at follow-up. |  |  |

Included papers did not allow sufficiently robust information to perform meta-analysis and so we present findings here narratively, with interventions grouped by: 1) Single-session interventions for emergency department crisis presentations; 2) Community-based crisis interventions (as i. exclusively in-home interventions, ii. interventions outside of the home but outside clinics, iii. exclusively clinic-based outpatient interventions, including intensive day treatment).

### Single-session interventions for emergency department crisis presentations

We found three papers which evaluated the effectiveness of single-session urgent response consultations in emergency departments. Two papers were service evaluations (Gillig, 2004; Parker et al., 2003), and one paper used a pre-intervention historical/retrospective control group (Wharff, Ginnis, & Ross, 2012). One paper was at critical risk of bias (Gillig, 2004), one at serious risk of bias (Parker et al., 2003) and one at low risk of bias (Wharff et al., 2012). Gillig (2004) investigated the effects of offering an emergency evaluation interview and a brief therapeutic intervention at a maximum of 24 hours after CYP presentation, using a supportive, reality-based and present-focused therapeutic approach. They reported that only 10% (n = 5) of the patients seen by the emergency consultation team were hospitalised right after the input was received, no patients were hospitalised in the month following the input and 4.2% (n = 2) patients were hospitalised 6 months later. Parker et al. (2003) analysed a Rapid Response Model (RRM) which offered consultations to CYP in acute mental health crises within 48 hours of their presentation to the ED, focusing on the crisis and risk. They used two sites: one site studied outcomes over four years (with pre-RRM, during RRM implementation, post-RRM termination and during RRM re-implementation as four time periods); another over two years (pre-RRM and post-RRM as two time periods). Findings were mixed. For the first site, no change was reported in the number of admissions over the four time periods but RRM stage had a significant effect on the monthly average length of inpatient stay: *F*(1, 42) = 3.1, *p* < .05). For the second site, there was a reported decrease in the percentage of admissions, with a reduction from 22% at pre-RRM to 2% at post-RRM (χ^2^ = 31.6, *N* = 340, d.f. = 1, *p* < .001). Wharff et al. (2012) investigated a single-session family-based crisis intervention (FBCI) for suicidal adolescents, delivered in a paediatric ED and focusing on constructing a safety plan and encouraging family communication. They reported a reduction in numbers admitted during the implementation of FBCI (55% to 35%, *p* < .0001).

### Community-based treatments

We found fourteen papers investigating the effectiveness of community-based treatments for CYP presenting in crisis. Nine were of home interventions, three were community interventions outside of the home but outside clinics and two were exclusively clinic-based outpatient interventions, including day treatment.

#### i) Exclusively in-home interventions

We found nine papers studying in home interventions – five studying multi-systemic therapy (MST), and four papers of other family-based interventions in the home.

Five papers investigated the effectiveness of multisystemic therapy (MST) delivered at home and which combined a range of therapeutic approaches. All five papers were RCTs, although four were in effect different outcomes from one single trial (Henggeler et al., 2003; Henggeler et al., 1999; Huey et al., 2004; Schoenwald, Ward, Henggeler, & Rowland, 2000). The risk of bias assessment revealed that four papers were at low risk of bias (Henggeler et al., 2003; Henggeler et al., 1999; Huey et al., 2004; Rowland et al., 2005; Schoenwald et al., 2000) while one raised some concerns (Rowland et al., 2005).

The effects of MST on admission rates and length of stay were investigated by two papers in two separate trials (Rowland et al., 2005; Schoenwald et al., 2000). In one RCT, in a sample of CYP in mental health crisis assessed to require an admission, MST was compared to hospitalization (Schoenwald et al., 2000). At the end of treatment, 44% of those assigned to the MST group (approximately 4 months post-referral), were admitted to hospital and mean length of stay was lower than the hospitalized control group (mean days 2.39 versus 8.82, *t* = 3.91, *p* = .001). In a second RCT, CYP at risk of out-of-home placement (e.g. inpatient hospitalization, group homes, foster care) were randomly assigned to receive either MST or usual care (Rowland et al., 2005). The study did not report on the number of patients hospitalised following the intervention in either group. However, number of days spent in out-of-home placement per month were lower in the intervention group (mean days 3.75 versus 11.83, *F* = 5.68, *p* = .025).

Potential psychological benefits of MST were investigated in four papers, three reporting outcomes from a single trial comparing MST to inpatients, and one from another trial comparing MST to usual care. Compared to hospitalisation (Henggeler et al., 1999; Huey et al., 2004), MST was reported to have superiority in improving a number of psychological measures. Post intervention changes were superior compared to admission controls for the caregiver rated externalising symptoms (*F*(1,102) = 6.55, *p* < .011), teacher rated externalising symptoms (*F*(1, 45) = 4.10, *p* < .048), youth reported family adaptability (*F*(2, 220) = 3.28, *p* = .039), caregiver rated family cohesion (*F*(2, 206) = 6.56, *p* < .001) and youth reported suicide attempts (*t* = 3.60, *p* < .001). However a further paper of 12-16 month follow-up revealed that generally superiority was not sustained (Henggeler et al., 2003). Change in self-esteem was initially superior in the hospitalized group (*F*(1, 109) = 7.72, *p* = .006), however again this was not sustained at 12–16-month follow-up. For MST compared to treatment as usual, (Rowland et al., 2005) there was superior change in youth-rated externalising symptoms (*F*(1, 25) = 4.62, *p* = .041) and internalising symptoms (*F*(1,28) = 6.05, *p* = .021) for the MST group.

We found four papers of other family-based interventions delivered at home which reported on psychological outcomes. One was an RCT (Wilmshurst, 2002), one was a study using a matched sample control group (Schmidt, Lay, Göpel, Naab, & Blanz, 2006) and two were uncontrolled studies investigating pre- to post-treatment changes (Lay, Blanz, & Schmidt, 2001; Mosier et al., 2001). Bias assessment revealed that two papers were at serious risk of bias (Lay et al., 2001; Mosier et al., 2001), one paper was at moderate risk (Schmidt et al., 2006) while the other paper raised some concerns (Wilmshurst, 2002). Neither paper described proportions of CYP receiving the alternative interventions who required admission In the RCT, Wilmshurst (2002) reported that the in-home treatment was superior to a 5-day residential programme in improving internalising symptoms at follow-up (*F* (2,62) = 3.92, *p* = .025). Comparing the intervention to a matched control group receiving hospitalisation, Schmidt et al. (2006) reported that by the end of treatment, hospitalisation was superior at improving psychological symptoms related to major DSM-IV diagnoses (*p* < .001), child-rated behaviour (*p* = .02), parent-rated behaviour (*p* = .03), and increased functioning in more domains (family, peers, interests, and autonomy) than the in-home treatment (family, interests, autonomy). Lay et al. (2001) compared pre- and post-treatment outcomes in a sample of CYP requiring an intervention as an alternative to admission presenting with high-risk externalising behaviours. Improvements were reported in the total symptoms related to major DSM-IV diagnoses (*p* < .001) and child-rated psychosocial functioning (*p* < .001). Mosier et al. (2001) also compared pre- and post-treatment outcomes of an in-home famility therapy for CYP requiring restrictive and intensive mental health treatment. In comparison to pre-treatment, post-treatment improvements were reported in clinical symptoms for a number of CYP, with some being reported to have recovered. No statistical tests were conducted for this comparison. The end-of-treatment mean clinical symptoms of the group was compared to the normative scores of patients receiving inpatient/outpatient treatment, but no significant differences were reported.

#### ii) Interventions outside of the home but outside of hospital clinics

We found three papers which evaluated the effectiveness of community interventions outside of the home but away from clinic settings. The stated aim of these interventions was to stabilize the patients by offering an array of services, delivered flexibly in the community depending on need (e.g., schools). One paper was an RCT (Winsberg, Bialer, Kupietz, Botti, & Balka, 1980) and two papers were service evaluations (Darwish, Salmon, Ahuja, & Steed, 2006; Vanderploeg, Lu, Marshall, & Stevens, 2016). The risk of bias assessment revealed that one paper was at high risk of bias (Winsberg et al., 1980), one was at critical risk (Vanderploeg et al., 2016) and one was at serious risk (Darwish et al., 2006). Only one of these papers, Darwish et al. (2006), reported on admission outcomes for their intervention, reporting that their community intensive therapy decreased admissions compared to a pre-intervention historical group (decreasing to 1 person per year over 5 years versus 6 people per year at pre-implementation, no statistics presented). Two papers reported on psychological outcomes. Winsberg et al. (1980) reported pre and post intervention scales and hospitalization on a group of CYP in crisis, on a range of psychological parameters. Post-intervention, the community treatment improved aggressivity symptoms (*t* = 2.58, *p* < .05), inattentiveness (*t* = 3.53, *p* < .01), hyperactivity (*t* = 4.27, *p* < .01), school behaviour (*t* = 2.58, *p* < .05), and overall child behaviour (*t* = 2.22, *p* < .05). In comparison, hospitalisation improved only inattentiveness (*t* = 2.32 *p* < .05). There was however no comparison of differences between the interventions and hospitalized groups. Vanderploeg et al. (2016) studied changes in a sample of CYP receiving emergency mobile psychiatric services however they did not compare their outcomes to a control group. They reported that improvements were achieved in parent-rated problem severity (*t* = -8.53, *p* < .001), clinician-rated problem severity (*t* = -21.24, *p* < .001) parent-rated child functioning (*t* = 4.70, *p* < .001) and clinician-rated child functioning (*t* = 14.96, *p* < .001).

#### iii) Exclusively clinic-based interventions, including intensive day treatment

We found four papers which evaluated the effects of interventions in clinic-based settings (one including intensive day treatment) in CYP presenting in crisis as alternatives to hospitalization. One paper was an RCT (Silberstein, Mandell, Dalack, & Cooper, 1968), one paper used a matched sample control group (Greenfield, Hechtman, & Tremblay, 1995) and two papers were uncontrolled pre-post intervention studies (Asarnow, Berk, Hughes, & Anderson, 2015; Kiser et al., 1996). The risk of bias assessment revealed that one paper was at moderate risk of bias (Asarnow et al., 2015), one at high risk of bias (Silberstein et al., 1968), one at serious risk of bias (Greenfield et al., 1995) and one at critical risk (Kiser et al., 1996).

Silberstein et al. (1968) compared 4 separate groups of CYP presenting with psychosis receiving: 1) parental counselling with or 2) without medication; 3) no counselling plus medication; and 4) no counselling plus placebo. There were no differences in rates of admissions between groups. Greenfield et al. (1995) investigated the outcomes of implementing an emergency room follow-up team, and reported that in comparison to a period before its implementation, the admission rate of CYP presenting to the emergency room in psychiatric crisis decreased by 16% (*p* < .001). No statistical difference was reported between the two groups in the number of hospitalizations occuring per patient after a second emergency room visit.

Kiser et al. (1996) reported on psychological outcomes of an outpatient day program for CYP in mental health crisis as an alternative to admission. Post-intervention, improvements were reported in being withdrawn (parent report: *t* = 5.25, *p* < .001; CYP report : *t* = 2.91, *p* = .005), somatic complaints (parent report: *t* = 3.11, *p* = .003; CYP report: *t* = 2.76, *p* = .008), anxious/depressed (parent report: *t* = 3.95, *p* < .001; CYP report: *t* = 3.95, *p* < .001), social problems (parent report: *t* = 2.70, *p* = .008; CYP report: *t* = 2.38, *p* = .021), thought problems (parent report: *t* = 5.66, *p* < .001; CYP report: *t* = 3.28, *p* = .002), attention problems (parent report: *t* = 4.28, *p* < .001; CYP report: *t* = 2.89, *p* = .06), delinquent behaviour (parent report: *t* = 32.49, *p* = .015; CYP report: *t* = 3.77, *p* < .001), aggressive behaviour (parent report: *t* = 4.49, *p* < .001; CYP report *t* = 3.59, *p* < .001); sex problems (significant for CYP report only: *t* = 4.66, *p* < .001), total problems (parent report: *t* = 6.24, *p* < .001; CYP report *t* = 5.58, *p* < .001), internalising (parent report: *t* = 4.90, *p* < .001; CYP report: *t* = 4.94, *p* < .001) and externalising (parent report: *t* = 4.84, *p* < .001; CYP report *t* = 4.90, *p* < .001). Both parents and CYP also reported that at follow-up, family functioning was improved in the following domains: roles (parent report only: *t* = 2.68, *p* = .009), affective involvement (CYP report only: *t* = 2.24, *p* = .03) and behaviour control (parent report: *t* = 3.41, *p* = .001; CYP report: *t* = 3.48, *p* = .001). Follow-up improvements were also reported in rates of school suspensions (from 39% to 36%, χ^2^ = 8.78, df = 1, p < .01), CYP being a good-quality friend (from 65% to 88%, χ^2^ = 6.45, df =1, p < .05), incarceration rates (from 4.7% to 6.3%, χ^2^ = 19.6, df = 1, p < .01) and trouble with the police (from 13.4% to 16.4%, χ^2^ = 11.6, df = 1, p < .01). Asarnow et al. (2015) reported on psychological outcomes of an outpatient intervention (the SAFETY program) delivered to adolescent suicide attempters. They reported pre- to post-treatment improvements in all outcomes measured: suicide attempts (*t* = 2.42, *p* = .019, *d* = .64), active suicide behaviour and ideation (*t* = 2.63, *p* = .019, *d* = .59), passive suicide ideation (*t* = 2.56, *p* = .016, *d* = .39), total suicidality score (*t* = 2.70, *p* = .011, *d* = .46), CYP reported youth depression symptoms (*t* = 4.53, *p* < .001, *d* = .91), parent-reported parental depression symptoms (*t* = 3.47, *p* = .002, *d* = .71), hopelessness (*t* = 5.58, *p* < .001, *d* = 1.01), social adjustment total score (*t* = 6.13, *p* < .001, *d* = 1.27), social adjustment at school (*t* = 3.53, *p* = .002, *d* = .90), social adjustment with peers (*t* = 5.36, *p* < .001, *d* = 1.11), social adjustment with the family (*t* = 2.79, *p* = .009, *d* = .66) and social adjustment in the spare time (*t* = 2.76, *p* = .01, *d* = .52).

## Discussion

In this systematic review of studies of alternatives to inpatient admissions for CYP presenting with a mental health crisis, we found a range of published studies on interventions in different settings. We found studies describing interventions in emergency departments, the home, other community settings and hospital-based clinics. In general, the level of evidence was poor with less than half of included studies RCTs, of which only half were considered of low risk of bias in bias assessments. Studies also varied with regard to consistency of reporting on measures on preventing admissions and psychological outcomes. This meant that robust data for meta-analysis was insufficient. The greatest level of evidence came from home treatments, in particular MST. MST was reported as improving a range of psychological parameters associated with risk for CYP (such as suicide attempts) and benefits for families (adaptation and cohesion though not maintained at 4 months); and though a large proportion of CYP appeared to still ultimately be admitted (in one study 44%), there was evidence that length of stay from these admissions was reduced compared to admission alone. We found some evidence suggesting that brief emergency department-based interventions could have a beneficial impact on admission rates, but none of these studies were RCTs, and there was no information on impact upon psychological parameters in any paper. Evidence for other community interventions, and clinic-based interventions were scarce, and generally of low quality. However, we found some evidence for reduction in admission rates and improvements in post-intervention symptom severity, child and family functioning, although these were not compared to outcomes of control groups.

Our review did not find sufficient amount of quality data to recommend a specific type of intervention for CYP presenting in crisis, a similar conclusion to the two other systematic reviews on this topic which included searches from over 6 years ago (Kwok et al., 2016; Shepperd et al., 2009). However, the evidence we have presented provides useful information for the development of new and existing services, including the potential for mix-models of care, or “menus” of care for individual patients’ needs by understanding variable benefits of different models. Given the challenges associated with the complexity of such CYP presenting in crisis, especially with regard to risk, the limited availability of good quality data is perhaps understandable. However, with such presentations increasing, and pressure on inpatient units rising (Children’s Commissioner, 2020), this is clearly an area which needs to see an increase in research as a priority. With new emphasis on improvement for CYP with mental health disorders, especially those presenting in crisis (Ougrin et al., 2018), it is likely that new models will develop. It is important that as they do so, they are robustly evaluated, in particular with comparison to controls (including for example pre-intervention controls), with consistent measurement and reporting of success at reducing absolute numbers of admissions, duration of admissions and also psychological impacts for CYP and families.

Studies should also report detail on change between groups of intervention and control, for a large proportion of studies we found in our review presented only pre and post values for intervention and control separately, and this impedes the opportunity for an appropriate pooling of studies in meta-analysis (Higgins, Churchill, Chandler, & Cumpston, 2017).

Beyond the limitations which we have highlighted above, our review has a number of strengths. We used an a priori search strategy of multiple databases, with defined inclusion and exclusion criteria for the studies and two independent researchers performed searches. We also investigated on a large range of intervention types by also including non-RCTs, in comparison to previous reviews that looked at RCTs only. We reported on all outcomes described and had two independent researchers to conduct thorough bias assessments, with a third providing final adjucation.

In conclusion, although we found a range of interventons in different settings, the quality of studies was insufficient to allow for an overall recommendation. Interventions using multi- systemic therapy at home had the best quality, with evidence suggesting benefits around avoiding admissions, length of admission and psychological outcomes. However, these interventions generally failed to show long-term effects. New models of care should be robustly evaluated using consistent outcomes.

## Data Availability

This is a systematic review, we are willing to share studies found.

## Appendix A

Search terms used for all databses: ((children and adolescents OR children OR adolescents OR youth) AND mental health AND (crisis OR crises)) AND (emergency department OR a&e OR ‘alternatives to hospital admission’ OR ‘home treatment’ OR ‘community based crisis’ OR ‘alternative care’ OR ‘short stay hospital’ OR ‘acute day hospital’ OR ‘acute ward’ OR ‘crisis houses’ OR ‘family based treatment’ OR ‘multisystemic therapy’ OR ‘crash pad*’ NOT (homeless OR homelessness))

## Appendix B

### Results of risk of bias assessment for randomised-controlled trials using the ROB2 tool

| Author (year, country) | Randomisation process | Notes | Deviations from intended interventions | Notes | Missing outcome data | Notes | Measurements of the outcome | Notes | Selection of the reported results | Notes | Overall bias |
| --- | --- | --- | --- | --- | --- | --- | --- | --- | --- | --- | --- |
| Henggeler et al (1999), USA | Low | No significant baselines differences reported between the two groups. Participants were randomly assigned to groups. | Low | No deviations mentioned | Low | No significant missing data reported | Low | Used validated outcome measures | Low | Results for all measured were reported, regardless to their statistical significance. | Low |
| Henggeler et al (2003), USA | Low | No significant baselines differences reported between the two groups. Participants were randomly assigned to groups. | Low | No deviations mentioned | Low | No significant missing data reported | Low | Used validated outcome measures | Low | Results for all measured were reported, regardless to their statistical significance. | Low |
| Huey et al (2003), USA | Low | No significant baselines differences reported between the two groups. Participants were randomly assigned to groups. | Low | No deviations mentioned | Low | No significant missing data reported | Low | Used validated outcome measures | Low | Results for all measured were reported, regardless to their statistical significance. | Low |
| Rowland et al (2005), USA | Some concerns | Baseline differences were found for child delinquency with the treatment group scoring higher than the usual services group | Low | No deviations mentioned | Some concerns | Follow-up data only available for 31 out of the 55 people that initially consented to take part in the study. No reason was given for why this happened for the majority of the patients whose data was not available | Low | Used validated outcome measures | Low | Results for all measured were reported, regardless to their statistical significance | Some concerns |
| Schoenwald et al (2000), USA | No information | No statistical tests were conducted to investigate baseline group differences although descriptive statistics were reported. Participants were randomly assigned to groups | Low | No deviations mentioned | Low | No significant missing data reported | Low | Used validated outcome measures | Low | Results for all measured were reported, regardless to their statistical significance. | Low |
| Silberstein, Mandell, Dalack, Cooper & Island (1967), USA | No information | No statistical tests were conducted to investigate baseline group differences although descriptive statistics were reported. Participants were randomly assigned to groups | Some concerns | The intervention plan was not described in detail and the paper did not mention if treatment protocol was adhered to | Low | No significant missing data reported | High | Test-retest reliability and validity of some of the measures used not mentioned | Low | Results for all measured were reported, regardless to their statistical significance. | High |
| Wilmshurt (2002), USA | Low | No statistical tests were conducted to investigate baseline group differences although descriptive statistics were reported. Participants were randomly assigned to groups | Low | No deviations mentioned | Some concerns | The paper mentions that a high proportion of teacher-rated and youth-rated data is missing. Although a clear explanation was given for the missingness of teacher data, no explanation was given for the missingness of youth-rated data. | Low | Used validated outcome measures | Low | Results for all measured were reported, regardless to their statistical significance. | Some concerns |
| Winsberg et al. (1980), USA | Low | No significant baselines differences reported between the two groups. Participants were randomly assigned to groups. | Low | No deviations mentioned | Low | No significant missing data reported | Low | Used validate outcome measures | High | Results on 2 of the subscales of the Behaviour Rating Scale were not presented | High |

### Results of risk of bias assessment for non-randomised trials using the ROBINS-I tool

| Reference | Confounding | Notes | Selection of participants | Notes | Classification of interventions | Notes | Deviations from intended intervention | Notes | Missing data | Notes | Measurement of outcome | Notes |
| --- | --- | --- | --- | --- | --- | --- | --- | --- | --- | --- | --- | --- |
| Asarnow, Berk, Hughes & Anderson (2015), USA | Moderate | Single group study but standardization used to control for possible confounders | Moderate | Some pre-established eligibility criteria used | Not Applicable | Single group design | Low | No deviations mentioned | Moderate | Although substantial missing data was reported, the authors used an appropriate statistical analysis to account for it | Low | Validated and reliable outcome measures used |
| Darwish, Salmon, Ahuja, & Steed (2006), UK | Critical | Retrospective control group and no statistical analysis conducted to determine baseline group differences | Serious | No pre-established eligibility criteria used. | Moderate | Retrospective study of different time periods in which patients attending a clinic either received or did not receive the new intervention | Low | No deviations mentioned | Low | No significant missing data reported | Low | Validated and reliable outcome measures used |
| Gilig (2004), USA | Critical | Single group design - potential for confounding | Serious | No pre-established eligibility criteria used. | Not applicable | Single group design | Low | No deviations mentioned | Low | No significant missing data report | Low | Validated and reliable outcome measures used |
| Greenfield, Hechtman, & Tremblay | Critical | Retrospective control group and no statistical analysis conducted to determine | Serious | No pre-established eligibility criteria used. | Moderate | Retrospective study of different time periods in which | Low | No deviations mentioned | Low | No significant missing data reported | Low | Validated and reliable outcome |
| (1995), Canada |  | baseline group differences |  |  |  | patients attending a clinic either received or did not receive the new intervention |  |  |  |  |  | measures used |
| Kiser et al (1996), USA | Critical | Single group design - potential for confounding | Serious | No pre-established eligibility criteria used. | Not applicable | Single group design | Low | No deviations mentioned | Low | No significant missing data reported | Low | Validated and reliable outcome measures used |
| Lay, Blanz & Schmidt (2001), Germany | Critical | Single group design - potential for confounding | Low | Clear pre-established eligibility criteria used. | Not applicable | Single group design | Low | No deviations mentioned | Low | No significant missing data reported | Low | Validated and reliable outcome measures used |
| Mosier, Burlingame & Wells (2001), USA | Serious | Single group design but statistical analysis conducted to determine baseline group differences in comparison to normative scores on the same scale for a sample receiving inpatient/outpatient treatment | Serious | No pre-established eligibility criteria used. | Not applicable | Single group design | Low | No deviations mentioned | Moderate | Although substantial missing data was reported, the authors used an appropriate statistical analysis to account for it | Low | Validated and reliable outcome measures used |
| Parker et al (2003), Canada | Critical | Retrospective study - potential for confounding | Serious | No pre-established eligibility criteria used. | Moderate | Retrospective study of different time periods in which patients attending a clinic either received or did not receive the new intervention | Low | No deviations mentioned | Low | No significant missing data reported | Low | Validated and reliable outcome measures used |
| Schmidt, Lay, Gopel, Naab, & Blanz (2006), Germany | Moderate | Although matched samples were used in the trial, the authors could not recruit the same number of patients in the control group as the experimental condition. | Low | Clear pre-established eligibility criteria used. | Low | Treatment and control groups used | Low | No deviations mentioned | Low | No significant missing data reported | Low | Validated and reliable outcome measures used |
| Vanderploeg, Lu, Marshall & Stevens (2016), USA | Critical | Single group design - potential for confounding | Serious | No pre-established eligibility criteria used. | Not applicable | Single group design | Low | No deviations mentioned | Low | No clear information on whether there was any missing data or not | Low | Validated and reliable outcome measures used |
| Wharff, Ginnis & Ross (2012) USA | Low | Matched control group used, and statistical analysis were conducted to determine differences between groups at baseline. No differences were found. | Low | Clear pre-established eligibility criteria used. | Moderate | Matched retrospective comparison group | Low | No deviations mentioned | Low | No significant missing data reported | Low | Validated and reliable outcome measures used |

## References

Alderwick, H., & Dixon, J. (2019). The NHS long term plan. In: British Medical Journal Publishing Group.

Asarnow, J. R., Berk, M., Hughes, J. L., & Anderson, N. L. (2015). The SAFETY Program: a treatment-development trial of a cognitive-behavioral family treatment for adolescent suicide attempters. Journal of Clinical Child & Adolescent Psychology, 44(1), 194–203.

Commissioner, C. s. (2020). Who are they? Where are they? 2020. Children in tier 4 mental health units. Technical report. Retrieved from

Darwish, A., Salmon, G., Ahuja, A., & Steed, L. (2006). The community intensive therapy team: Development and philosophy of a new service. Clinical Child Psychology and Psychiatry, 11(4), 591–605.

Gillig, P. M. (2004). Child & adolescent psychiatry: An adolescent crisis service in a rural area. Psychiatric Services, 55(12), 1363–1365.

Green, J., Jacobs, B., Beecham, J., Dunn, G., Kroll, L., Tobias, C., & Briskman, J. (2007). Inpatient treatment in child and adolescent psychiatry–a prospective study of health gain and costs. Journal of child psychology and psychiatry, 48(12), 1259–1267.

Greenfield, B., Hechtman, L., & Tremblay, C. (1995). Short-term efficacy of interventions by a youth crisis team. The Canadian Journal of Psychiatry, 40(6), 320–324.

Hawton, K., Bergen, H., Kapur, N., Cooper, J., Steeg, S., Ness, J., & Waters, K. (2012). Repetition of self-harm and suicide following self-harm in children and adolescents: Findings from the Multicentre Study of Self-harm in England. Journal of child psychology and psychiatry, 53(12), 1212–1219.

Henggeler, S. W., Rowland, M. D., Halliday-Boykins, C., Sheidow, A. J., Ward, D. M., Randall, J., … Edwards, J. (2003). One-year follow-up of multisystemic therapy as an alternative to the hospitalization of youths in psychiatric crisis. Journal of the American Academy of Child & Adolescent Psychiatry, 42(5), 543–551.

Henggeler, S. W., Rowland, M. D., Randall, J., Ward, D. M., Pickrel, S. G., Cunningham, P. B., … Hand, L. D. (1999). Home-based multisystemic therapy as an alternative to the hospitalization of youths in psychiatric crisis: Clinical outcomes. Journal of the American Academy of Child & Adolescent Psychiatry, 38(11), 1331–1339.

Higgins, J., Churchill, R., Chandler, J., & Cumpston, M. (2017). Cochrane handbook for systematic reviews of interventions version 5.2. 0 (updated February 2017), Cochrane, 2017. Available from Cochrane Community.

Huey, S. J., Henggeler, S. W., Rowland, M. D., Halliday-Boykins, C. A., Cunningham, P. B., Pickrel, S. G., & Edwards, J. (2004). Multisystemic therapy effects on attempted suicide by youths presenting psychiatric emergencies. Journal of the American Academy of Child & Adolescent Psychiatry, 43(2), 183–190.

Jennings, S., & Child, C. A. P. B. Healthy London Partnership–Children and Young People Programme: Improving care for children and young people in mental health crisis in London: Recommendations for transformation of services.

Kiser, L. J., Millsap, P. A., Hickerson, S., Heston, J. D., Nunn, W., Pruitt, D. B., & Rohr, M. (1996). Results of treatment one year later: Child and adolescent partial hospitalization. Journal of the American Academy of Child & Adolescent Psychiatry, 35(1), 81–90.

Kovess-Masfety, V., Husky, M. M., Keyes, K., Hamilton, A., Pez, O., Bitfoi, A., … Otten, R. (2016). Comparing the prevalence of mental health problems in children 6–11 across Europe. Social psychiatry and psychiatric epidemiology, 51(8), 1093–1103.

Kwok, K. H. R., Yuan, S. N. V., & Ougrin, D. (2016). Alternatives to inpatient care for children and adolescents with mental health disorders. Child and Adolescent Mental Health, 21(1), 3–10.

Lancet, T. (2020). Child mental health services in England: a continuing crisis. Lancet (London, England), 395(10222), 389.

Lay, B., Blanz, B., & Schmidt, M. H. (2001). Effectiveness of home treatment in children and adolescents with externalizing psychiatric disorders. European child & adolescent psychiatry, 10(1), S80–S90.

Mahajan, P., Alpern, E. R., Grupp-Phelan, J., Chamberlain, J., Dong, L., Holubkov, R., … Sonnett, M. (2009). Epidemiology of psychiatric-related visits to emergency departments in a multicenter collaborative research pediatric network. Pediatric emergency care, 25(11), 715–720.

Merikangas, K. R., He, J.-p., Burstein, M., Swanson, S. A., Avenevoli, S., Cui, L., … Swendsen, J. (2010). Lifetime prevalence of mental disorders in US adolescents: results from the National Comorbidity Survey Replication–Adolescent Supplement (NCS-A). Journal of the American Academy of Child & Adolescent Psychiatry, 49(10), 980–989.

Miller, D. A., Ronis, S. T., Slaunwhite, A. K., Audas, R., Richard, J., Tilleczek, K., & Zhang, M. (2020). Longitudinal examination of youth readmission to mental health inpatient units. Child and Adolescent Mental Health, 25(4), 238–248.

Morgan, C., Webb, R. T., Carr, M. J., Kontopantelis, E., Green, J., Chew-Graham, C. A., … Ashcroft, D. M. (2017). Incidence, clinical management, and mortality risk following self harm among children and adolescents: cohort study in primary care. Bmj, 359.

Mosier, J., Burlingame, G. M., Wells, M. G., Ferre, R., Latkowski, M., Johansen, J., … Walton, E. (2001). In-home, family-centered psychiatric treatment for high-risk children and youth. Children’s Services: Social Policy, Research, and Practice, 4(2), 51–68.

Newton, A. S., Ali, S., Johnson, D. W., Haines, C., Rosychuk, R. J., Keaschuk, R. A., … Klassen, T. P. (2009). A 4-year review of pediatric mental health emergencies in Alberta. Canadian Journal of Emergency Medicine, 11(5), 447–454.

O’Herlihy, A., Worrall, A., Lelliott, P., Jaffa, T., Hill, P., & Banerjee, S. (2003). Distribution and characteristics of in-patient child and adolescent mental health services in England and Wales. The British Journal of Psychiatry, 183(6), 547–551.

Ougrin, D., Corrigall, R., Poole, J., Zundel, T., Sarhane, M., Slater, V., … Heslin, M. (2018). Comparison of effectiveness and cost-effectiveness of an intensive community supported discharge service versus treatment as usual for adolescents with psychiatric emergencies: a randomised controlled trial. The Lancet Psychiatry, 5(6), 477–485.

Paranjothy, S., Evans, A., Bandyopadhyay, A., Fone, D., Schofield, B., John, A., … Long, S. J. (2018). Risk of emergency hospital admission in children associated with mental disorders and alcohol misuse in the household: an electronic birth cohort study. The Lancet Public Health, 3(6), e279–e288.

Parker, K. C., Roberts, N., Williams, C., Benjamin, M., Cripps, L., & Woogh, C. (2003). Urgent adolescent psychiatric consultation: from the accident and emergency department to inpatient adolescent psychiatry. Journal of adolescence, 26(3), 283–293.

Pittsenbarger, Z. E., & Mannix, R. (2014). Trends in pediatric visits to the emergency department for psychiatric illnesses. Academic Emergency Medicine, 21(1), 25–30.

Polanczyk, G. V., Salum, G. A., Sugaya, L. S., Caye, A., & Rohde, L. A. (2015). Annual Research Review: A meta-analysis of the worldwide prevalence of mental disorders in children and adolescents. Journal of child psychology and psychiatry, 56(3), 345–365.

Rowland, M. D., Halliday-Boykins, C. A., Henggeler, S. W., Cunningham, P. B., Lee, T. G., Kruesi, M. J., & Shapiro, S. B. (2005). A randomized trial of multisystemic therapy with Hawaii’s Felix Class youths. Journal of Emotional and Behavioral Disorders, 13(1), 13–23.

Schmidt, M. H., Lay, B., Göpel, C., Naab, S., & Blanz, B. (2006). Home treatment for children and adolescents with psychiatric disorders. European child & adolescent psychiatry, 15(5), 265–276.

Schoenwald, S. K., Ward, D. M., Henggeler, S. W., & Rowland, M. D. (2000). Multisystemic therapy versus hospitalization for crisis stabilization of youth: Placement outcomes 4 months postreferral. Mental Health Services Research, 2(1), 3–12.

Shepperd, S., Doll, H., Gowers, S., James, A., Fazel, M., Fitzpatrick, R., & Pollock, J. (2009). Alternatives to inpatient mental health care for children and young people. Cochrane Database of Systematic Reviews(2).

Silberstein, R. M., Mandell, W., Dalack, J. D., & Cooper, A. (1968). Avoiding institutionalization of psychotic children. Archives of General Psychiatry, 19(1), 17–21.

Sterne, J. A., Hernán, M. A., Reeves, B. C., Savović, J., Berkman, N. D., Viswanathan, M., … Boutron, I. (2016). ROBINS-I: a tool for assessing risk of bias in non-randomised studies of interventions. Bmj, 355.

Sterne, J. A., Savović, J., Page, M. J., Elbers, R. G., Blencowe, N. S., Boutron, I., … Eldridge, S. M. (2019). RoB 2: a revised tool for assessing risk of bias in randomised trials. Bmj, 366.

Torio, C. M., Encinosa, W., Berdahl, T., McCormick, M. C., & Simpson, L. A. (2015). Annual report on health care for children and youth in the United States: national estimates of cost, utilization and expenditures for children with mental health conditions. Academic pediatrics, 15(1), 19–35.

Vanderploeg, J. J., Lu, J. J., Marshall, T. M., & Stevens, K. (2016). Mobile crisis services for children and families: advancing a community-based model in Connecticut. Children and youth services review, 71, 103–109.

Vizard, T., Sadler, K., & Ford, T. (2020). Mental Health of Children and Young People in England, 2020: Wave 1 follow up to the 2017 survey. NHS Digital: https://digital.nhs.uk/data-and-information/publications/statistical/mental-health-of-children-and-young-people-in-england/2020-wave-1-follow-up.

Wasserman, D., Cheng, Q., & Jiang, G.-X. (2005). Global suicide rates among young people aged 15-19. World psychiatry, 4(2), 114.

Wharff, E. A., Ginnis, K. M., & Ross, A. M. (2012). Family-based crisis intervention with suicidal adolescents in the emergency room: a pilot study. Social work, 57(2), 133–143.

Wilmshurst, L. A. (2002). Treatment programs for youth with emotional and behavioral disorders: An outcome study of two alternate approaches. Mental Health Services Research, 4(2), 85–96.

Winsberg, B. G., Bialer, I., Kupietz, S., Botti, E., & Balka, E. B. (1980). Home vs hospital care of children with behavior disorders: a controlled investigation. Archives of General Psychiatry, 37(4), 413–418.

Worrall, A., O’Herlihy, A., Banerjee, S., Jaffa, T., Lelliott, P., Hill, P., … Brook, H. (2004). Inappropriate admission of young people with mental disorder to adult psychiatric wards and paediatric wards: cross sectional study of six months’ activity. Bmj, 328(7444), 867.

